# SuReCAN: a suite of user-friendly Galaxy machine learning workflows to predict survival and treatment response of cancer patients

**DOI:** 10.1101/2025.08.12.25333156

**Authors:** Jie Ju, Daria Koppes, Andrew P. Stubbs, Yunlei Li

**Affiliations:** Erasmus Medical Center

## Abstract

Cancer is one of the leading lethal causes worldwide, with enormous impact on healthcare, economy and society. One of the main challenges of clinical treatment planning is that patients usually have diverse clinical outcomes given the same diagnosis and treatments. To enable personalized cancer therapeutic planning, (bio)medical data analyses using machine learning (ML) models are introduced to efficiently extract informative biological patterns from the massive volume of complex biological data, aiding in cancer patients’ stratifications.

For biomedical researchers without computational biology background, the gap between clinical practice and computational approaches is prominent and hinders the usage of machine learning in medical research. To fill this gap, we created a collection of ML workflows on the Galaxy platform named SUrvival and REsponse prediction for CANcer patients (SuReCAN) for clinicians and biologists to build and deploy predictive ML classifiers. Being freely available and accessible, SuReCAN automates the data analysis process and enables the clinicians and researchers to perform a broad range of predictive tasks. It contains a toolkit of three ML modules with various existing and newly implemented methods on Galaxy: A data normalization module, a feature selection module, and an ML classifier module.

We exhibited the utility of SuReCAN with a few real-world datasets to identify pancreatic ductal adenocarcinoma (PDAC) patients’ survival-correlated subtypes and to predict drug response outcomes based on various omics data from patient tumor samples. As a result, all workflows achieved a median accuracy of over 0.8 in PDAC survival-correlated subtype classification. In particular, the workflow combining the feature selection method “SVM-based *RFECV*” and the Support Vector Machine classifier consistently outperformed the other workflows, while all classifiers have shown their superiority on different omics data.

Importantly, SuReCAN is not only applicable for the clinical prediction tasks shown in the test cases but also suitable for new classifier development and deployment with clinical observations provided by the users. Providing a collection of user-friendly ML workflows, SuReCAN stratifies patients based on their biomedical profiling in a data-driven way and assists biomedical researchers with clinical decision-making and scientific discoveries.

## Background

Cancer is the second leading cause of death worldwide, which means one out of six death cases is due to cancer^1^. Unfortunately, the aging population leads to a continuously increased number of cancer-related deaths nowadays. The heterogeneity of the tumors is one of the leading determinates of cancer therapeutic defects and poor prognosis of cancer patients^2^; the diversity of cell subgroups largely influences the cancer progression and treatment response, and results in distinct survival and drug response outcomes in patients^3^. Therefore, personalized treatment planning for cancer patients is urgent.

The development of technologies enables the measurements of a large volume of biological data at the molecular level, such as omics, which significantly accelerates the exploration of biomarkers for prognosis prediction and drug discovery. In the meanwhile, to handle the fast-growing data effectively, interdisciplinary studies together with artificial intelligence have become an integral part of modern medicine. Machine learning (ML) models have been utilized in the medical field to make accurate predictions for each patient’s clinical outcome and assist the clinical decision-making^4^. Classification is an ML technique that stratifies samples into target classes. In the medical field, for example, classification models have been applied to the molecular profile of individuals to distinguish the responders from non-responders of a therapeutic plan^5,6^ or to stratify patients based on their predicted survival outcome^6,7^. Based on these predictions, surgical and therapeutic decisions could be made accordingly to maximize the benefits and minimize the side effects in patients.

The implementation and compiling of the classification tools for cancer research is a complex and technically intense process, requiring a solid understanding of different algorithms as well as the medical fields. Specifically, the construction of the ML classification models includes data normalization, feature selection, and classifier construction with parameter tuning. In each step, it is crucial to apply suitable methods for specific research questions. To bridge the computational and medical fields and make the classification tools amenable for medical researchers, there is a necessity to provide a user-friendly workflow that gives accurate outcome predictions directly based on the input omics data from the laboratories.

In this study, we introduce a collection of seven end-to-end prediction workflows based on omics data, named SUrvival, and REsponse prediction for CANcer patients (SuReCAN) which is implemented on the Galaxy platform^8^. The workflows are ready to be used for classifier construction and deployment in medical research, and are suitable for people without expertise in bioinformatics or ML. Specifically, SuReCAN contains a toolkit to cover the full process of building and deploying prediction models including data normalization, feature selection, and ML classifiers construction and training. A few built-in tools on the Galaxy platform were utilized in the workflows, and some new methods were implemented to complete the workflows and serve a broad range of classification purposes. We rigorously tested our workflows under three experimental settings of treatment response and survival-correlated subtype prediction on five omics datasets of pancreatic ductal adenocarcinoma (PDAC) patients. In these experiments, the newly implemented and existing feature selection methods and classification models were evaluated and compared on the Galaxy platform. The workflows will provide clinicians and biologists with an efficient and effective toolkit to get patients’ clinical outcome predictions timely, helping clinical treatment planning and cancer research in a data-driven way.

## Workflow

SuReCAN completes the ML workflows with three connected modules, including data normalization, feature selection, and ML classifiers (**Figure 1**). In each module, different methods are either adopted directly from existing tools or implemented as new components on the Galaxy platform. Therefore, SuReCAN, as a collection of seven ML workflows, consists of combinations of different methods from data normalization, feature selection and machine learning modules to train and deploy classifiers for different research interests and predictive tasks. Additionally, the output of the feature selection module could be extracted as the predictors of the ML classifiers and become candidate biomarkers in the laboratories for further investigations and validations.

**Figure 1.**
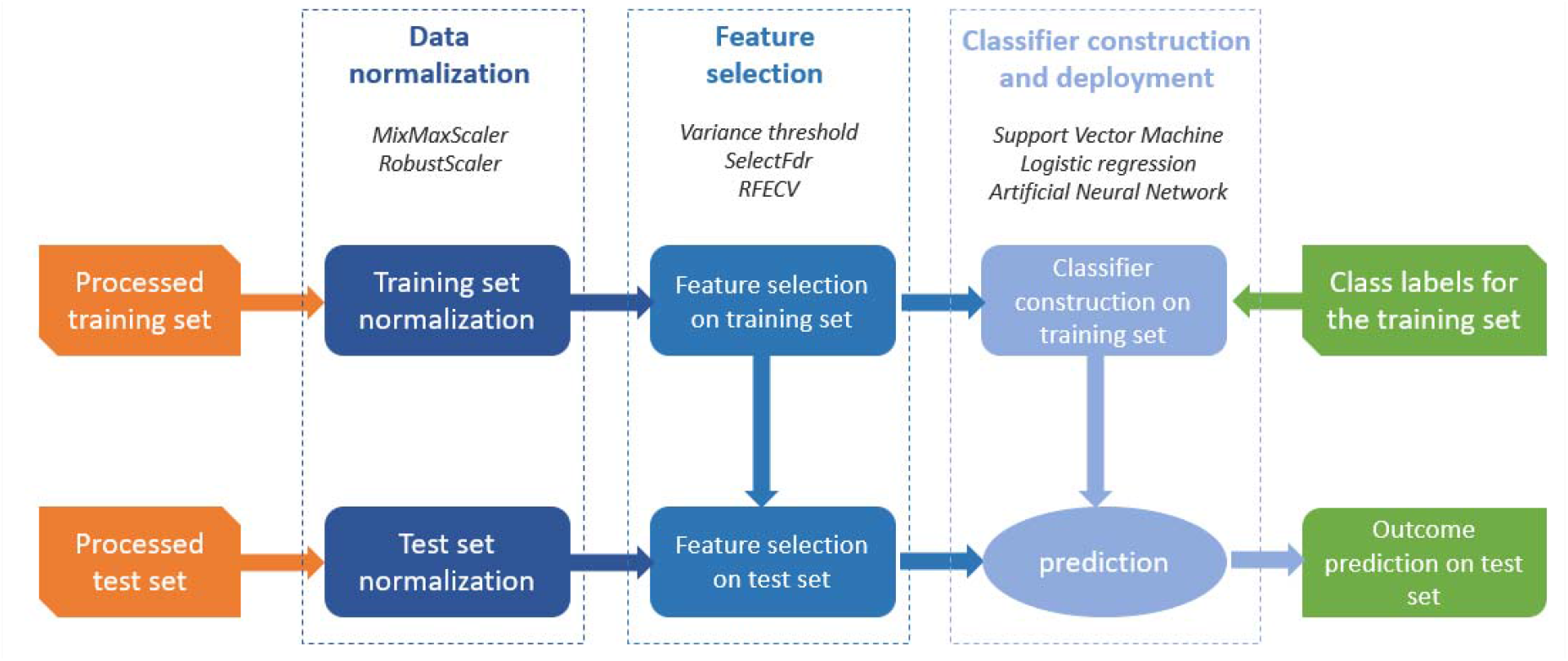
Schematic representation of SuReCAN machine learning workflow implemented in Galaxy.

SuReCAN takes in the omics data that has been properly pre-processed and stored in CSV files, with omics features as columns and samples as rows. They are often readily available as the output of the profiling experiment. The pre-processing methods should be determined based on the characteristics of the data^9^ and the research questions. For example, RNA-seq data need to be pre-processed into counts and normalized with methods like Transcripts Per Million (TPM) or fragments per kilobase of transcript per million fragments mapped (FPKM) to correct for the technical impact that potentially disrupts the biological effects of interest. Given the input files, the workflows start with performing an additional normalization to ensure that the measurements of the features are in the same range and adjust them to the same scale within the dataset. The normalization methods included in the workflows are MinMaxScaler and RobustScaler^10^. The MinMaxScaler linearly scales the feature values between 0 and 1, while the RobustScaler shifts the median values to zero and scales according to the quantile ranges of the features. The exact formula are shown in **Table 1**.

**Table 1.**
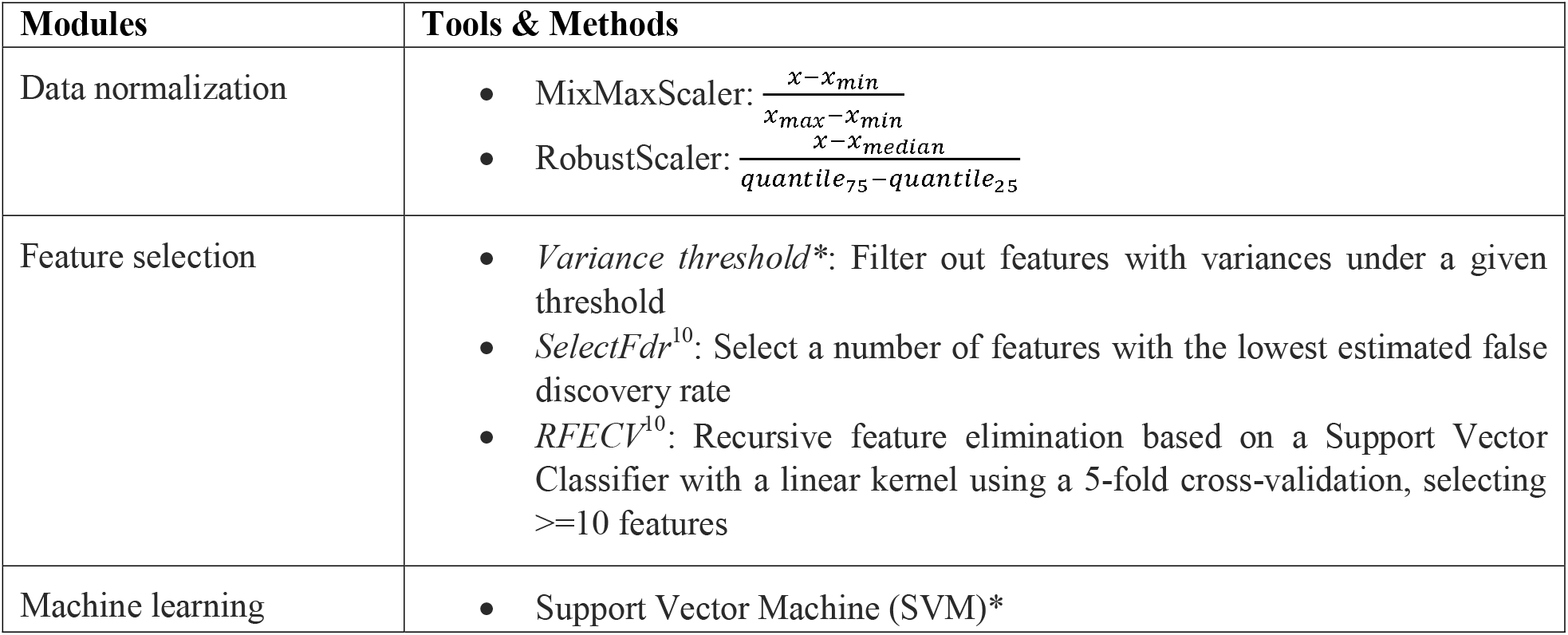
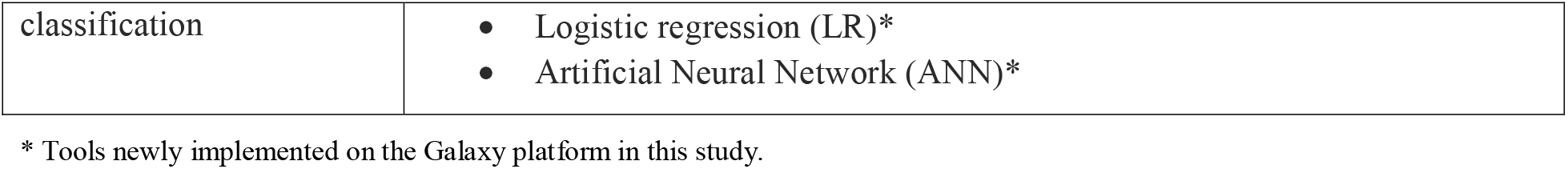
SuReCAN toolkit.

The next step is the selection of predictive features in order to reduce dimensionality and remove noise for the classification models. Three widely used methods are included: (1) The *variance threshold* is an unsupervised approach, which filters features with variances below a threshold given by the users. (2) The *selectFdr* is a supervised learning approach using statistical models, selecting features with the most significant correlations to the class labels according to the false discovery rate (FDR). (3) The *recursive feature elimination with cross-validation* (RFECV) is a supervised feature selection method, which filters out the least contributing features in a Support Vector Machine (SVM) model using a 5-fold cross-validation (CV) ^10^. The predictors identified in the feature selection step could be obtained from the module to understand the decision-making process in the predictive models.

The ML classifiers are constructed based on the selected features from the training set, and the predicted labels of the test sets are produced as the output of the workflows. The classifiers implemented in the workflow include Logistic Regression (LR), Support Vector Machine (SVM), and Artificial Neural Network (ANN). LR is a simple and robust classifier which uses a logistic function to model the probability of a binary outcome based on the input features. SVM, which is a powerful prediction model for complex datasets, aims to search for a hyper-plane that maximizes the separations between the data points in different classes in a high-dimensional feature space^11^. ANN is a deep learning model that enables non-linear interactions between the features, while a large amount of training samples is required for this method^12^. Hyper-parameters could be fine-tuned on the training sets to ensure the robustness of the classifiers.

## Implementation

All tools and their dependencies are installed on the Galaxy platform. The code is publicly available on the Github page (https://github.com/ErasmusMC-Bioinformatics/SuReCAN). The overview of SuReCAN toolkit is shown in **Table 1**. Besides the tools provided on the Galaxy platforms already, several newly-added methods were developed as below.

The *variance threshold* method for feature selection was realized using the *Table Compute* tool in Galaxy, in which the threshold of the sample/feature variances could be given by the users.

The SVM and LR models were built using the “Pipeline Builder” tool in Galaxy, with the “sklearn.svm” and “sklearn.linear” modules, respectively. The output of the “Pipeline Builder” tool could be used for hyperparameter optimization with the “Hyperparameter Search” tool with “GridsearchCV” or “RandomizedsearchCV”. The ANN architectures were constructed using *Keras* library and subsequently tuned with the tool “Hyperparameter Search” in Galaxy. Parameters and the range of grid searches were provided in Supplementary Table 1-3.

### Training Materials

The links to the workflows created in this study are listed in **Table 2**. A more detailed description of the usage of the entire workflow is given in our training manual on the Galaxy Training materials repository (https://github.com/ErasmusMC-Bioinformatics/SuReCAN). The workflow collection SuReCAN is freely available from Galaxy.

**Table 2.**
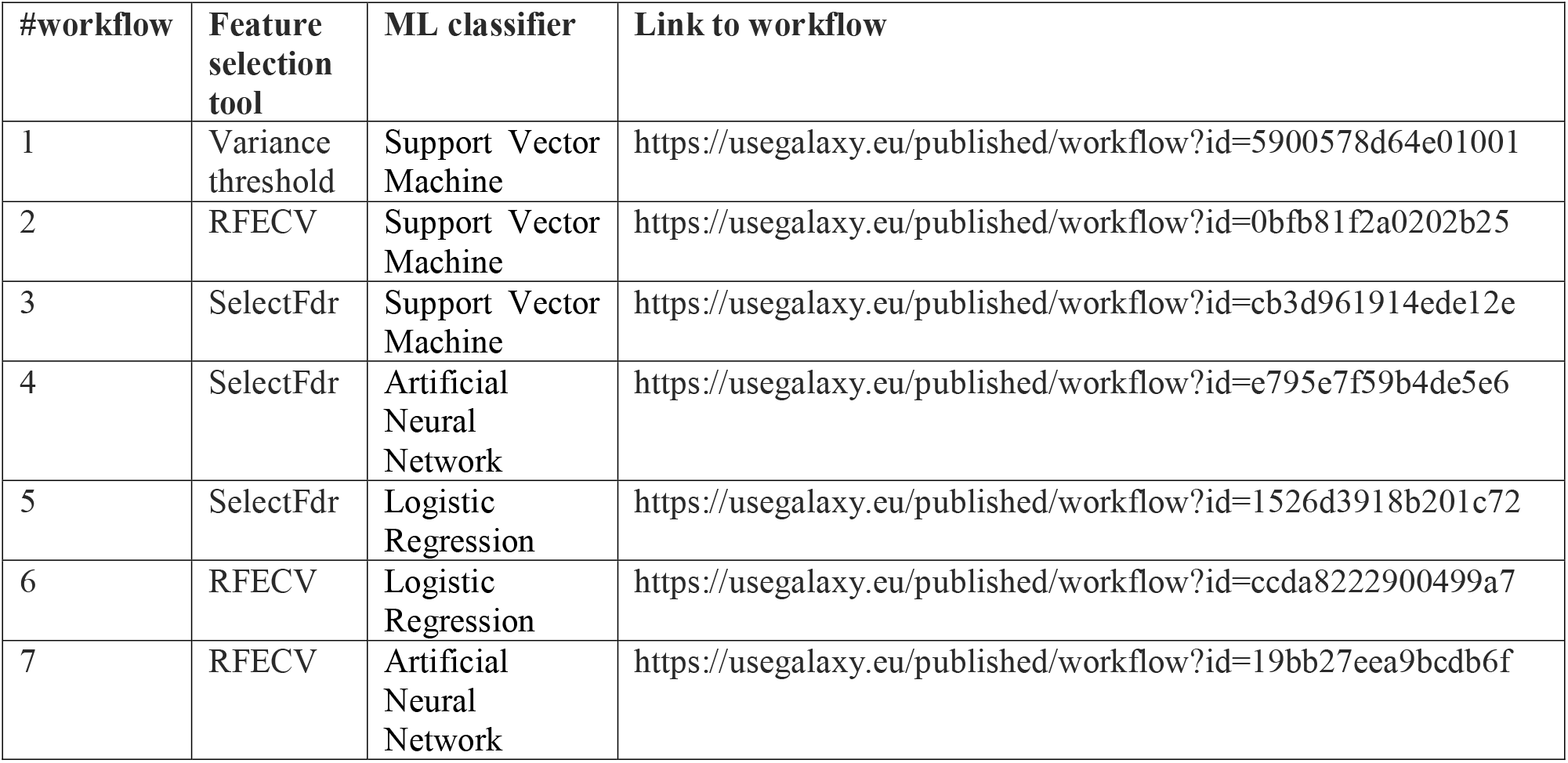
Links to the workflows in SuReCAN.

## Worked example: Survival and treatment response prediction

As a demonstration and validation of SuReCAN workflows on Galaxy, we utilized five real-world omics datasets of pancreatic cancer patients and performed three experiments to stratify the patients based on their survival-correlated subtypes and to predict their responses to a first-line chemotherapy. The workflows were stored and could be deployed directly for the prediction tasks of new observations. More importantly, the workflows could be used to develop customized classifiers suitable for the predictions of other research questions with the datasets generated and provided by the users.

### Background

Pancreatic cancer is one of the leading causes of cancer-related death worldwide, with a five-year survival rate of less than 10%^13^. Pancreatic ductal adenocarcinoma (PDAC), as the most aggressive subtype, accounts for 85% of pancreatic cancer^14^. The high heterogeneity of PDAC leads to distinct survival outcomes and responses to treatments in patients, which poses challenges to treatment planning. Specifically, only 20% of PDAC patients benefit from surgical resection, while the rest have a high risk of suffering from surgical complications^15^. Furthermore, currently PDAC patients in an advanced stage receive the first-line chemotherapy, consisting of FOLFIRINOX and gemcitabine, with a response rate of only 31.6% and 9.4%, respectively^16^, while the non-responders still have to go through the toxic procedure. Therefore, there is a necessity to predict the survivorship and responses to the therapies for PDAC patients and plan personalized treatments accordingly.

We aim to fill in the gap computationally, utilizing SuReCAN ML workflows constructed in Galaxy for these prediction tasks. In total, four omics datasets obtained from public databases were deployed in three experiments, to predict the PDAC patients’ survival-correlated subtypes and chemotherapy responses based on various types of omics data. Additionally, the performance of different feature selection and ML methods were assessed and compared.

### Datasets

The datasets used to test the SuReCAN ML workflows in Galaxy are listed in **Table 3**. Datasets 1-3 contain methylation array, mRNA-sequencing (mRNA-seq), and microarray data of primary PDAC tissues obtained from The Cancer Genome Atlas Program (TCGA) database^17^. The preprocessing of the datasets and prediction targets (i.e. survival-correlated PDAC subtype: aggressive/moderate) were described in our previous study ^18^. Dataset 4 contains mRNA sequencing data from PDAC tissues treated by chemotherapies (i.e. gemcitabine or FOLFIRINOX) from COMPASS study, which utilized RNA-seq data to identify predictive signatures for advanced PDAC ^19^. The PDAC patients’ response outcomes of chemotherapy treatment were evaluated according to RECIST criteria^20^ and aligned with clinical practices, stratifying the patients into disease control (including complete response, partial response, stable disease) and progressive disease. The gene features in dataset 4 with zero values in more than 40% of the samples were removed, followed by the same preprocessing as conducted in datasets 1-3.

**Table 3.** Descriptions of the five PDAC datasets used in this study.

| # | Data source | Data type | Class labels | #Samples | Sample origin |
| --- | --- | --- | --- | --- | --- |
| 1 | TCGA: PDAC | DNA methylation array | Aggressive/Moderate | 146 | Primary tumor samples |
| 2 | TCGA: PDAC | mRNA sequencing | Aggressive/Moderate | 146 | Primary tumor samples |
| 3 | TCGA: PDAC | microRNA sequencing | Aggressive/Moderate | 146 | Primary tumor samples |
| 4 | EGAS00001002543<br>* | mRNA sequencing | Disease control/Progressive disease | 157 | Locally advanced/metastatic tumor samples treated by chemotherapies |

PDAC: pancreatic ductal adenocarcinoma; TCGA: The Cancer Genome Atlas Program; \* Controlled dataset

### Experiments

We carried out three experiments to assess the feasibility and utility of the ML workflows in Galaxy. The respective training and test datasets (described in **Table 3**), feature selection methods, and classification models used in the three experiments are specified in **Table 4**.

**Table 4.**
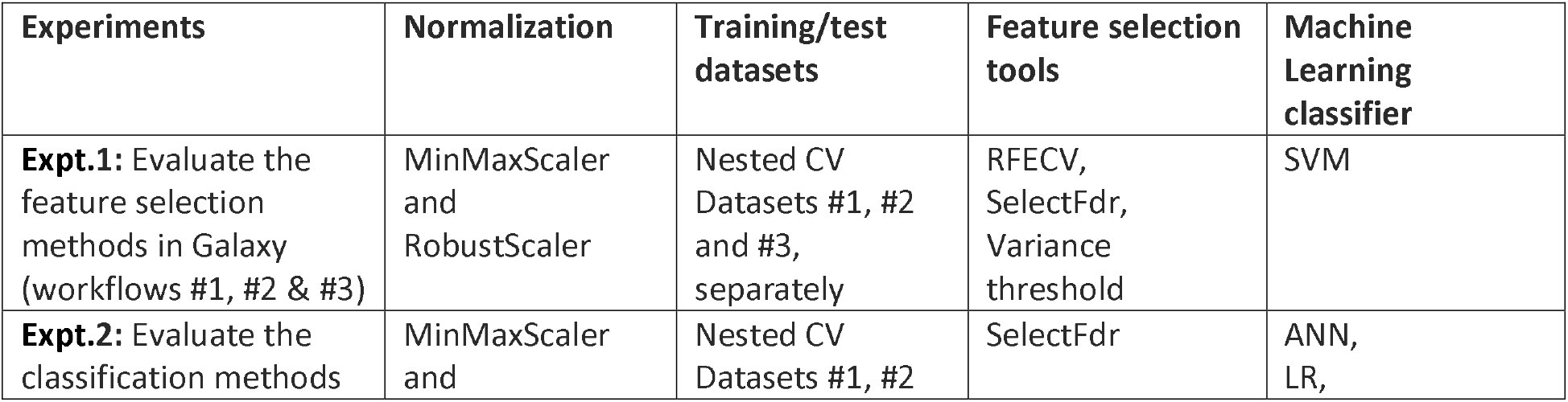
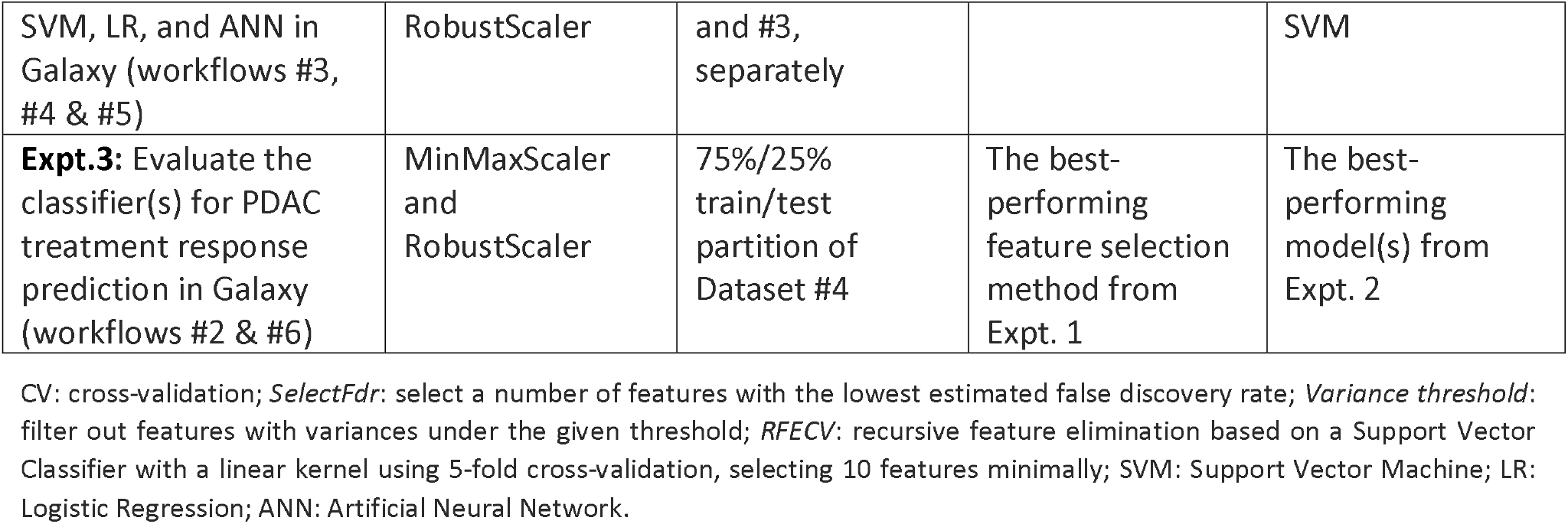
Overview of the three experiments as the worked examples of SuReCAN machine learning workflows.

Expts.1 & 2 aim to predict survival-correlated PDAC subtypes with nested cross-validation (CV) on datasets 1-3. In the nested CV, a 3-fold CV inner loop was constructed for the parameter tuning, and a 5-fold CV outer loop was performed for the performance evaluation of the models. The nested CV process was repeated 5 times and resulted in 25 estimations per experiment. More importantly, Expt.1 compared the performance of the feature selection methods *RFECV, SelectFdr*, and *variance threshold* based on the SVM classifier whereas Expt.2 compared classifiers ANN, LR, and SVM based on *SelectFdr* feature selection. All the comparisons between the experiments were carried out using the same data splits and tested by the Wilcoxon signed-rank test^21^ to assess the significance of the performance superiority.

For Expt.3, we aim to predict the chemotherapy treatment response in PDAC patients on datasets 4, using the best-performing feature selection method and ML classifier in Expts.1 & 2. In this experiment, dataset 4 was split into training and test sets with the partition of 75:25. Due to the class imbalance of the prediction targets, i.e. there were many more disease control patients than progressive disease patients in the datasets, the model performance was evaluated by three metrics, precision, recall, and F1-score.

### Results

In Expt.1, the SVM model was trained and validated with three feature selection methods *RFECV, selectFdr*, and *Variance threshold* on datasets 1-3. (**Figure 2A**). The results demonstrated a considerably high performance of the models constructed by SuReCAN on Galaxy, with a median accuracy of over 0.8 in all experiments. Furthermore, the prediction accuracy of the SVM model together with SVM-based *RFECV* was consistently higher than the other two methods with significance. This demonstrates the advantage of utilizing the SVM-based feature selection method before an SVM classifier. Therefore, *RFECV* was used in Expt.3.

**Figure 2.**
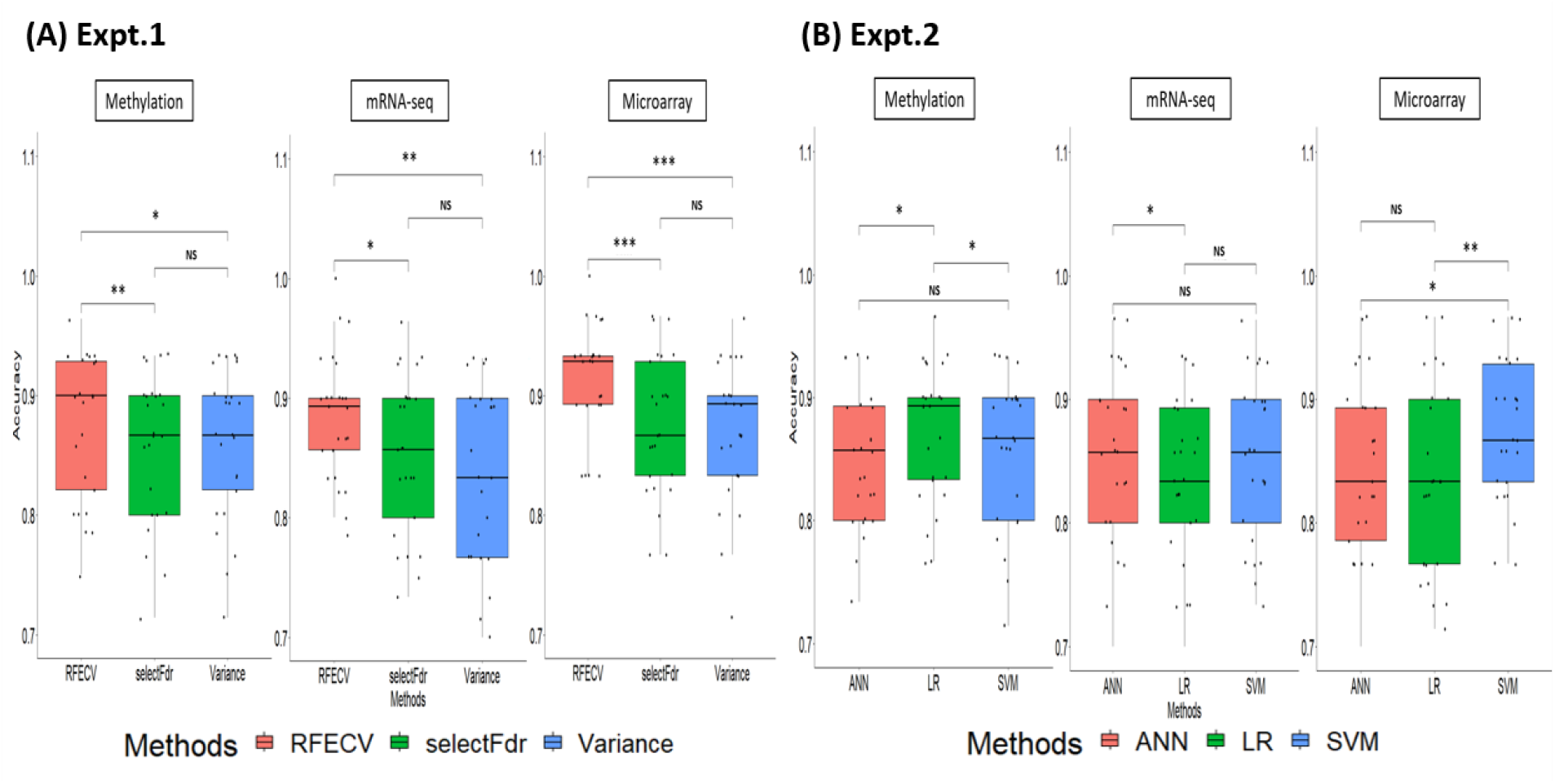
The resulting performance of SuReCAN for survival-correlated PDAC subtype prediction on different omics datasets (methylation, mRNA-seq, and microarray). **(A)** Expt. 1: The accuracy using different feature selection methods. **(B)** Expt. 2: The accuracy using different machine learning prediction models. The black horizontal line shows the median value of the accuracies. ns: not significant, *: p-value < 0.05, **: p-value < 0.01, ***: p-value < 0.001. All p-values were calculated using Wilcoxon signed-rank test.

In Expt.2, the performance of three ML classifiers (ANN, LR, and SVM) were compared on datasets 1-3 after *selectFdr* feature selection. Again, all the experiments achieved a median accuracy of over 0.8 (**Figure 2B**). The results showed that none of the classification models always outperformed the others, and all methods have shown their robustness in the prediction on different datasets. These results suggest the importance of implementing a broad range of classifier models and providing users with sufficient options for different research purposes.

Notably, the hyper-parameter searching was substantially faster in computation time for SVM and LR classifiers compared to ANN (i.e. SVM: 2min, LR:40s, ANN:2.5h, using the European Galaxy server). For this reason, only SVM and LR were used to test their applicability for the treatment response prediction for PDAC patients in Expt.3.

In Expt.3, PDAC patients’ responses to chemotherapy treatment were predicted using *RFECV* feature selection plus LR or SVM classifiers. Again, SVM achieved higher performance than LR in all performance evaluation metrics (**Figure 3**).

**Figure 3.**
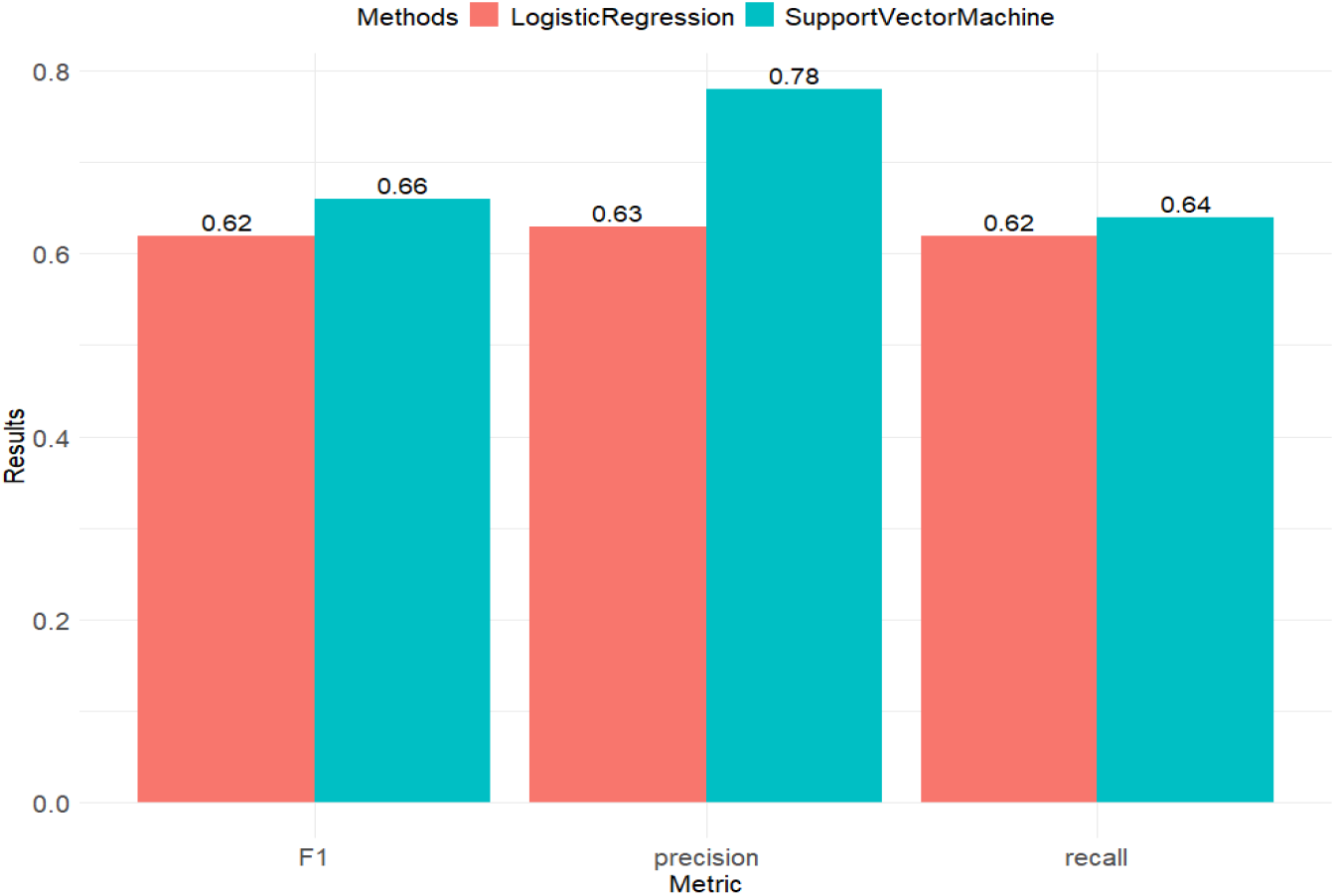
The performance of Logistic Regression and Support Vector Machine models on RNA dataset for chemotherapy treatment response prediction in PDAC patients.

## Discussions

In this work, we constructed and validated SuReCAN, a collection of ML workflows on the Galaxy platform. It is publicly available for any users such as clinicians and biologists to easily make predictions for patients’ clinical outcomes in a data-driven way and could be used to assist clinical decision-making and scientific research. The workflows contain three modules, including data normalization, feature selection, and ML classification. In addition to adopting the existing tools on Galaxy, new feature selection and machine learning methods were implemented in these modules to complete the workflows and serve a broader range of clinical needs and research interests.

We exhibited a real-world use case with four PDAC datasets for patients’ survival-correlated subtypes and treatment response prediction to extensively test and compare the performances of the existing and newly implemented methods in the workflows. The showcased experiments were based on public biomedical datasets and represent clinical research problems in practice. For the comparisons of feature selection and classifier methods in the prediction of PDAC patients’ survival-correlated subtypes, the results showed the superiority of SVM-based RFECV feature selection method when combined with SVM prediction model. Meanwhile, all classifier models exhibited their advantages on different omics, which demonstrated the importance of having a broad spectrum of choices in the workflow. However, when computation time is taken into account, simpler models such as LR and SVM exhibit advantages over complex models such as ANN. For the treatment response prediction task based on mRNA-seq data, we observed a superior performance by SVM classifier combined with RFECV feature selection. Therefore, we generally recommend using workflow #2 for these prediction tasks.

As to potential future extension of SuReCAN, evaluation metrics of the classification outcomes could be integrated into the workflows to make the model comparisons easier for users when constructing new classifiers. Furthermore, there are already tools on Galaxy for (bio)medical raw data preprocessing and quality check, which could be adapted and integrated into the workflows to automate the data engineering processes for the users. As a prevalent problem in medical datasets, class imbalance was also observed in our showcased studies due to the absence of non-responders receiving chemotherapies. Given the situation, adding algorithms overcoming the underrepresentation of the minority class in classifications would help improve the performance in the future.

Taken together, we demonstrate the utility of SuReCAN for the prediction tasks based on omics datasets. Besides the examples of the survival and treatment response of PDAC patients shown here, SuReCAN is also suitable for other similar prediction tasks, in other diseases, and using other molecular measurements. Meanwhile, the predictors selected for the predictions become candidate biomarkers for further experimental testing and validation. Furthermore, SuReCAN can be utilized to train new classifiers with clinical observations obtained and provided by users and make predictions for customized tasks, from which predictors could be identified. We believe that SuReCAN makes an important contribution in assisting medical research and clinical decision-making by bridging the medical research and machine learning fields with this representative collection of easy-to-use end-to-end workflows to analyze the big volume of (bio)medical data.

## Data Availability

All data produced in the present study are available upon reasonable request to the authors.

## Authors’ Contributions

YL conceived and supervised the study. DK implemented the SuReCAN workflow. JJ and DK conducted the analyses and wrote the manuscript. AP and YL reviewed the manuscript. All authors read and approved the final manuscript.

## Acknowledgement

This research was supported by an unrestricted grant of Stichting Hanarth Fonds, the Netherlands. This study was conducted with the support of the Ontario Institute for Cancer Research through funding provided by the Government of Ontario.

## Supplementary

**Supplementary table 1.** Support Vector Machine parameters.

| Parameters | Values |
| --- | --- |
| C | {0.0001, 0.0005, 0.001, 0.005, 0.01, 0.05, 0.1, 0.5} |
| Gamma | {0.001, 0.005, 0.01, 0.05, 0.1, 0.5, 1, 1.5, 2, 2.5, 3} |
| Coef0 | {0.001, 0.005, 0.05, 0.1, 0.25, 0.5, 0.75, 1} |

**Supplementary table 2.** Logistic Regression parameters.

| Parameters | Values |
| --- | --- |
| C | {0.0001, 0.0005, 0.001, 0.005, 0.01, 0.05, 0.1, 0.5, 1, 5, 10} |
| max iter | {100, 200, 300, 400, 500} |
| fit intercept | {True, False} |
| class weight | {None, balanced} |

**Supplementary table 3.** Artificial Neural Network parameters.

| Parameters | Values |
| --- | --- |
| loss function | binary cross-entropy |
| optimizer | stochastic gradient descent (SGD) |
| learning rate | {0.001, 0.01, 0.1} |
| momentum | {0.5, 0.9, 0.99} |
| batch size | {1, 2, 4, 8, 16, 32} |

